# Computational Modeling of Attentional Impairments in Disruptive Mood Dysregulation and Attention Deficit/Hyperactivity Disorder

**DOI:** 10.1101/2020.04.15.20063578

**Authors:** Simone P. Haller, Joel Stoddard, David Pagliaccio, Hong Bui, Caroline MacGillivray, Matt Joes, Melissa A. Brotman

## Abstract

**Objective:** Computational models provide information about cognitive components underlying behavior. When applied to psychopathology-relevant processes, they offer additional insight to observed differences in behavioral performance. Drift diffusion models have been successfully applied to investigate processing efficiency during binary choice tasks. Using these, we examine the association between psychopathology (irritability and inattention) and processing efficiency under different attentional demands.

**Method:** 187 youth with ADHD, DMDD, both disorders, or no major psychopathology (*M* age=13.09, *SD*=2.55; 34% female) completed an Eriksen Flanker task. Of these, 87 provided complete data on dimensional measures of the core symptom of DMDD, irritability, and those of ADHD, inattention and hyperactivity.

**Results:** In a categorical analysis (*n*=187), we found a ADHD-by-DMDD-by task condition interaction on processing efficiency (*b*=−1.07, *p*=.01, 95%CI=[−1.89,−.24]), where increases in processing efficiency for non-conflict conditions were larger in youth without psychopathology relative to patients. Analysis of symptom severity (*n*=87) across diagnosis was consistent with the above analysis, revealing an interaction between symptom dimensions and task condition on processing efficiency (b=−0.10, *p*=.018, 95%CI[−.18, −.02]). Inattention, and its combined effect with irritability, predicted the magnitude of difference in processing efficiency between conflict and non-conflict conditions.

**Discussion:** Reduced processing efficiency may represent a shared cognitive endophenotype between ADHD and DMDD. Youth high in irritability or inattention may have difficulty adjusting processing efficiency to changing task demands possibly reflecting impairments in cognitive flexibility.

## Introduction

Increased reaction time variability on cognitive tasks is among the most replicated behavioral alterations in ADHD (Huang-Pollock, Karalunas, Tam, & Moore, 2012; Kofler et al., 2013). Recent psychophysical theories suggest that such variability can be explained by processing inefficiency, which explains behavior that is not just more variable in speed but also slower and likely to result in errors (Huang-Pollock et al., 2017). However, reaction time variability may not be specific to ADHD but also observed in other psychiatric phenotypes (Kaiser et al., 2008). Particularly, very little work to date has examined reaction time variability in youth with severe, chronic irritability. Here, we provide evidence from both a categorical and continuous, transdiagnostic approach to demonstrate associations between irritability and attention deficits, reaction time variability, and processing efficiency.

In studying affective psychopathology, examining response time variability may yield important insights into relevant cognitive processing mechanisms in affective (Warren et al., 2020) and nonaffective contexts (Lawlor et al., 2019). The high rate of comorbidity between ADHD and DMDD as well as co-occurrence of irritability and inattention symptoms specifically in clinical populations (Althoff et al., 2016; Kircanski et al., 2017; Mayes, Waxmonsky, Calhoun, & Bixler, 2016; Pagliaccio et al., 2017), has impeded our examination of the relative contributions of irritability and attentional symptoms to reaction time variability. As noted above, response time variability is a stable feature of ADHD, present across diverse tasks (e.g., reward, cognitive control), with some evidence for moderation of group differences by task difficulty (Epstein et al., 2011). However, advances in computational modeling and clinical phenotyping allow us to examine the associations between irritability, ADHD symptoms, and reaction time variability.

Computational models measure latent cognitive constructs and can reveal mechanism-based psychopathology-related differences in behavioral performance. With regards to response time variability in ADHD, the drift diffusion model (DDM) has been particularly impactful (Karalunas, Huang-Pollock, & Nigg, 2012; Ziegler, Pedersen, Mowinckel, & Biele, 2016). The DDM accounts for reaction time, reaction time variability, and accuracy in fast, binary choice tasks (Ratcliff & McKoon, 2008). It is well-defined with strong theoretical underpinnings and excellent explanatory power for human behavior (Jones, 2017). In the DDM, latent constructs are represented by parameters coding the strength of evidence entering the decision process, the amount of accumulated evidence required to make a decision, as well as motor preparation and output to explain speed-accuracy trade-off effects (Ratcliff & Tuerlinckx, 2002). Because it accounts for reaction time variability while accounting for all behavior (accuracy and reaction time), the primary parameter of interest for the present study is the drift rate, *v*, generally interpreted as processing efficiency (i.e., a large value represents more rapid, less variable, and error-resistant responses).

In the current study, we compared children diagnosed with ADHD, DMDD, both, or no major psychopathology on their performance on an Erikson flanker task. A subsample of participants also provided dimensional measures of the core symptom of DMDD, irritability, and those of ADHD, inattention and hyperactivity, for a complementary transdiagnostic analysis of symptoms. Based on previous findings (Karalunas et al., 2012; Salum et al., 2014), we expect that youth with ADHD will have lower drift rates relative to healthy volunteers. The Flanker task includes attentional conditions which may facilitate processing or introduce interference; we expect interference to slow processing and therefore reduce drift rates (White, Ratcliff, & Starns, 2011). Thus, the attentional demands of the flanker task not only allow an examination of associations between irritability and ADHD symptoms and drift rate but also the change in these associations with changing attentional demands. This is first study to systematically investigate response time variability and processing efficiency in DMDD; thus, we are agnostic in our predictions about cognitive control impairments in DMDD only, as the contribution of comorbid ADHD to attention processing in this population remains unclear.

## Methods

### Participants

Two-hundred and twenty-one youth with and without psychopathology (healthy volunteers; HVs) participated in this study. Youth were recruited both locally and nationally through practitioner referrals and IRB approved advertisements. Written informed consent was obtained from parents and assent was obtained from children. The study was approved by the NIMH Institutional Review Board.

Diagnoses were made by master’s or doctoral level clinician trained to reliability (κ>0.9) using a modified Schedule for Affective Disorders and Schizophrenia for School-Age Children— Present and Lifetime Version (K-SADS-PL; Kaufman et al., 1997) with an additional module to assess presence of DMDD (Wiggins et al., 2016). For all participants, full scale IQ<70 (determined by the Wechsler Abbreviated Scale of Intelligence; WASI; Wechsler, 2011), history of head trauma, neurological disorder, pervasive developmental disorder, medical illness preventing study participation, cardinal bipolar symptoms, post-traumatic stress disorder, schizophrenia, schizophreniform disorder, schizoaffective illness, current major depressive disorder or substance abuse within 2 months were exclusionary.

A total of 34 youth were excluded, 32 for task performance (see Methods for criteria), one child was excluded as the model failed to produce valid parameter estimates and one child was identified as a high leverage value in the multilevel model (Cook’s distance >2.5). Characteristics of the remaining 187 participants are presented in Table 1. Age and Sex did not significantly differ between diagnostic groups. Significant differences in FSIQ emerged across groups.

**Table 1.** Sample characteristics by diagnostic group

|  | **HV** | **ADHD**<br>**only** | **DMDD**<br>**only** | **DMDD**<br>**+**<br>**ADHD** | **Test Statistic** | **P** | **Post-hocs<sup>a</sup>** |
| --- | --- | --- | --- | --- | --- | --- | --- |
| N | 47 | 44 | 19 | 77 |  |  |  |
| Age<br>M(SD)<br>yrs | 13.61<br>(2.55) | 12.95<br>(2.59) | 12.95<br>(2.90) | 12.88<br>(2.43) | $F(3,183)=0.90$ | .45 | |
| Sex<br>(% F) | 43 | 27 | 53 | 27 | $\chi^2(3)=6.93$ | .07 | |
| IQ M(SD) | 113 (15) | 115<br>(12) | 114<br>(10) | 108<br>(13) | $F(3,183)=3.09$ | .02 | ADHD, HV><br>DMDD+ADHD,<br>DMDD |
| Accuracy | 0.99<br>(0.02) | 0.98<br>(0.03) | 0.99<br>(0.02) | 0.98<br>(0.03) | $F(3,183)=2.28$ | .08 | |
| Mean RT<br>(correct) | 491.15<br>(80.60) | 503.45<br>(77.83) | 511.81<br>(73.21) | 509.91<br>(73.99) | $F(3,183)=0.66$ | .58 | |

*Note.* HV: healthy controls, ADHD: attention dysregulation/hyperactivity disorder, DMDD: disruptive mood dysregulation disorder
<sup>a</sup>Pairwise comparisons using t-tests with pooled SD at $p<.05$ .

A subsample of *n*=87 youth (30 youth with ADHD only, 7 youth with DMDD only, 38 youth with DMDD+ADHD and 12 HV youth) also completed the parent-reported Affective Reactivity Index (ARI; Stringaris et al., 2012) and the Conners’ Parent Rating Scale (CPRS – hyperactive and inattentive subscale, raw scores (Conners, Sitarenios, Parker, & Epstein, 1998; CPRS – hyperactive and inattentive subscale, raw scores), assessing irritability and ADHD severity, respectively.

### Task

Children were presented with a modified version of the Eriksen Flanker task that used arrows rather than letter stimuli (Erikson & Erikson, 1974; Scherer, 1994). Five stimuli were arrayed horizontally with a central target (a left or right pointing arrow) and two distractors on each side. Conditions were: 1) congruent: in which distractors were identical to the target; 2) incongruent: in which distractor arrows were pointed opposite to the target; and 3) neutral: in which distractors were squares. Each trial started with a fixation cross (500 ms) followed by stimuli and response window (1000 ms) and then a blank screen (1500 ms). Figure 1 depicts the stimuli and trial sequence. The task was presented as one continuous block of 130 trials, taking 6.5 minutes to complete. Condition order was pseudo-randomly determined to maintain the same frequency for each condition. Each participant experienced the same order of trial condition.

**Figure 1.**
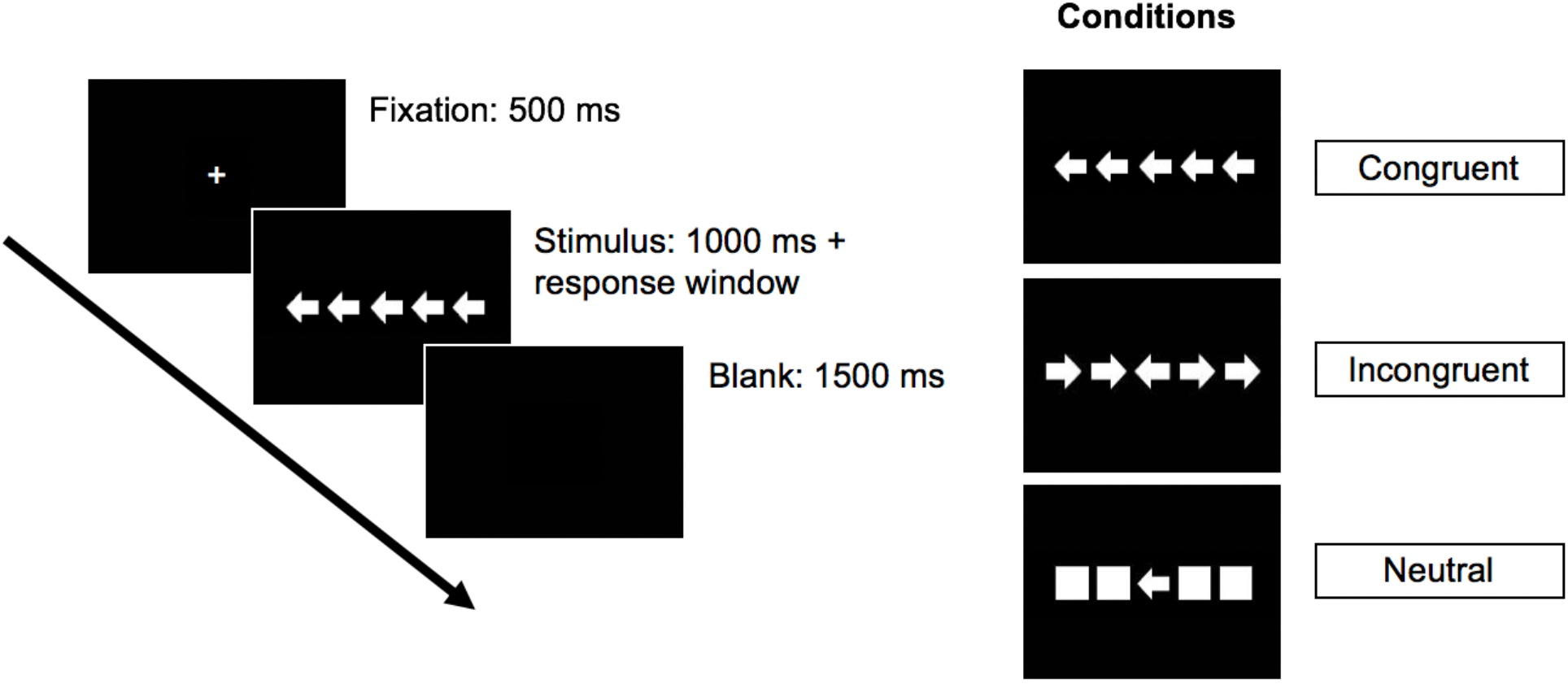
Trial sequence of the modified Eriksen Flanker task. Each trial started with a fixation cross followed by the stimulus presentation. A blank screen was presented between trials.

The task was presented via E-Prime 1.1 Build 1.1.4.1 or E-Prime 1.2 Build 1.2.1.847 on a laptop computer running Windows XP version 2002 or Windows 7 Professional. Stimuli arrays were 5.93×1.22 cm presented on a laptop placed at a viewing distance of approximately 60 cm. Participants responded to the target’s direction with their corresponding index finger, each placed on the laptop’s “a” (left) and “l” (right) keys. Children completed a practice task of 30 trials and were trained to 83% (25/30 correct responses) accuracy before proceeding to the task.

### Statistical analyses

#### Data preparation

Task data were prepared for analyses by removing non-physiologic anticipation responses (RT<150ms; 0.2% of all trials) and trials without responses. Participants who performed at or below chance (≤50% accuracy on any condition) or were too poorly engaged in the task (an overall non-response rate ≥15%) were excluded from further analyses (*n*=32). All analysis was performed in *R* Version 3.5 using the “lme4” package (Bates et al., 2015).

#### Drift Diffusion Model

As a speeded binary choice reaction time paradigm generally completed with high accuracy and speed, the Flanker task is fit for the application of the Drift Diffusion Model. Previous work has successfully used drift diffusion modeling in the analysis of Flanker task data (White et al., 2011). Diffusion parameters were estimated for each condition from all non-missing trials of the task using the full distribution of reaction times (i.e., congruent, incongruent and neural trials, errors). Diffusion parameters were estimated from the trial-by-trial data for each participant using the *fast-dm* modeling program Version 30.2 (Voss & Voss, 2007; Voss, Voss, & Lerche, 2015). For each condition, the model estimated parameters for a correct decision versus an error. Drift rate (*v*) varied across condition. Boundary separation (*a*), and non-decision time (*T*) were computed for each participant and constant across conditions. Bias, *zr*, was set to 0.5. The Maximum Likelihood approach was used to estimate fit.

To examine model performance, we compared simulated to the empirical data. We used the *construct-samples* tool, part of *fast-dm*, to simulate data sets from participants’ specific parameter set. For each participant, one dataset with *n*=1000 trials was simulated for each condition. Group reaction time distributions were constructed by averaging the quantiles of individual reaction time distributions. Given the small number of errors, medians were plotted rather than quantiles (White et al., 2011). Quantile probability plots (Ratcliff & Tuerlinckx, 2002) are a standard method to represent the distribution of reaction time by accuracy for all conditions in a task. See Figure 2 for a quantile probability plot for five reaction time quartiles (0.1, 0.3, 0.5, 0.7 and 0.9) for the empirical and simulated data for all youth.

**Figure 2.**
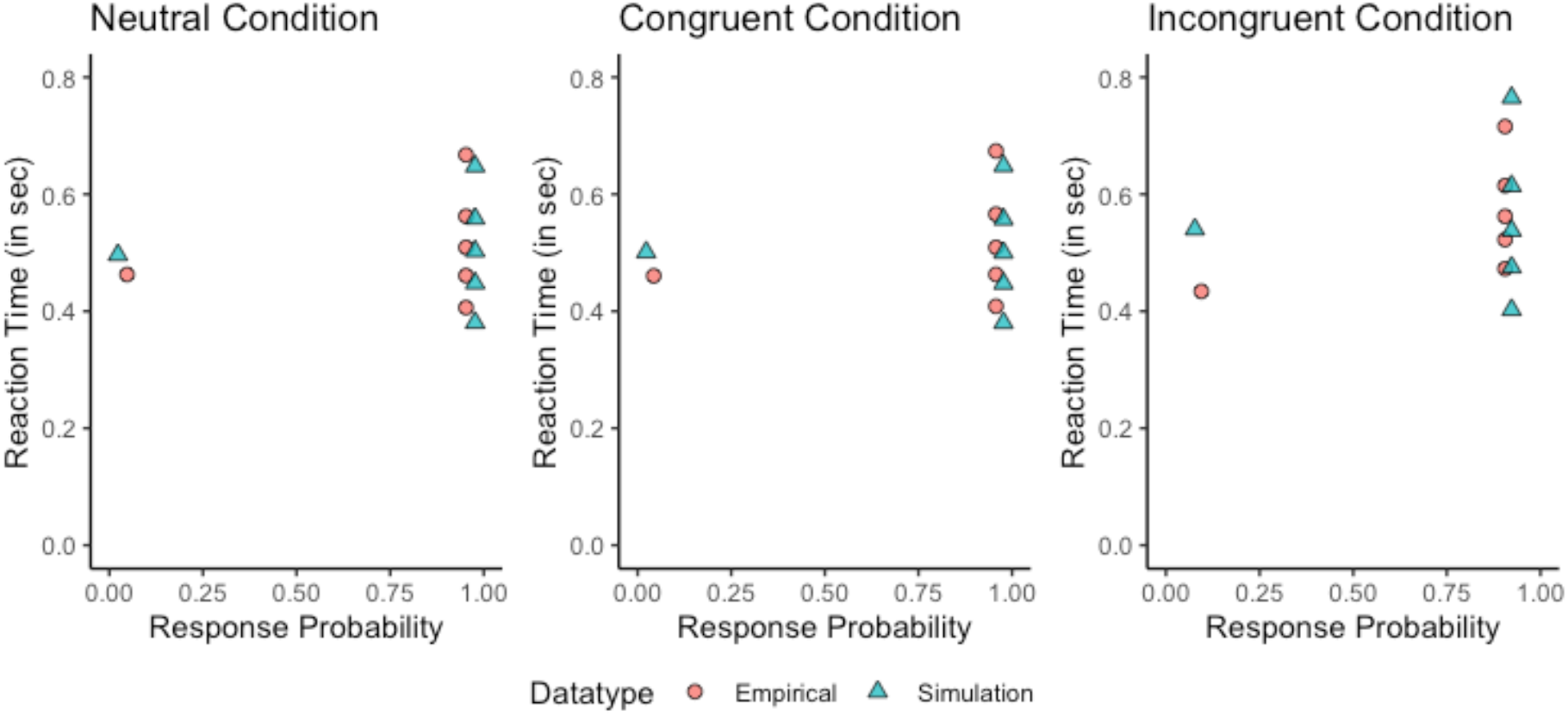
Quantile probability plot for pooled data from all youth. For each condition, the 0.1, 0.3, 0.5, 0.7, and 0.9 quantiles of reaction time are averaged across participants or simulations of their parameter set. Correct responses are on the right of each panel and represent almost all responses (average accuracy >92% by condition). Overall, the model reasonably represents behavior, especially in the neutral and congruent conditions. In the incongruent condition, the model underestimates the fastest 30% of reaction times, especially the fastest 10%. This is expected as attention dynamics that represent early interference effects on the drift rate are not represented with a constant drift rate (White et al., 2011).

A multilevel model testing the effects of diagnosis (DMDD and ADHD as separate predictors) and task condition (three level repeated measure) and task condition with Age and FSIQ as covariates and participant as a random effect was used to examine the DMM drift rate (v) parameter of interest. In a smaller sample (*n*=87), we used similar multilevel models to examined associations between continuous measures of irritability and ADHD (hyperactivity and inattentiveness tested separately) and task condition predicting drift rate with, task condition with Age and FSIQ as covariates. Because the ARI has a floor effect, it was converted to a binary factor representing low or high irritability symptoms. As including Sex in the models did not affect results, the models are presented without this additional covariate.

Supplementary Materials include analyses examining reaction time variability (intra-subject variability of reaction time measured as the coefficient of variation (CoV), the standard deviation/mean RT for correct responses by condition). We also include analyses exploring the relationship between CoV and drift rate for each condition using Pearson correlations.

## Results

### Diffusion Model Parameters

As expected response time variability as measured by CoV was associated with drift rate all *r*s(185)>-.62, p<.001, for each task condition. See Supplementary Materials for an analysis of CoV and a correlation matrix between drift rate and CoV by condition (Table S2).

### Diagnostic Associations

For the multivariate categorical analysis of the DDM drift rate parameter (*v*), a significant DMDD-by-ADHD-by-task condition interaction emerged (*n*=187; *b*=−1.07, *p*=0.01, 95%CI=[−1.89, −0.24], see Table 2 for full model). Differences emerged such that all patient groups showed lower drift rates compared to youth without psychopathology between non-conflict (neutral and congruent) and conflict conditions (see Figure 3).

**Table 2.** Mixed model regression table <u>fo</u>r dri<u>f</u>t rate predicted <u>b</u>y diagnostic classification.

| *Predictors* | *Estimates* | **v** |  |
| --- | --- | --- | --- |
|  |  | *CI* | *p* |
| Intercept | -4.15 | -7.15 – -1.14 | **0.007** |
| ADHD | -0.31 | -1.08 – 0.46 | 0.430 |
| DMDD | -0.75 | -1.75 – 0.25 | 0.144 |
| Condition [Congruent] | -0.22 | -0.58 – 0.14 | 0.240 |
| Condition [Incongruent] | -2.53 | -2.89 – -2.17 | **<0.001** |
| Age | 0.42 | 0.32 – 0.52 | **<0.001** |
| FSIQ | 0.04 | 0.02 – 0.06 | **<0.001** |
| ADHD * DMDD | 0.71 | -0.52 – 1.93 | 0.258 |
| ADHD * Condition [Congruent] | 0.18 | -0.34 – 0.70 | 0.507 |
| ADHD * Condition [Incongruent] | 0.62 | 0.10 – 1.14 | **0.021** |
| DMDD * Condition [Congruent] | 0.07 | -0.60 – 0.75 | 0.829 |
| DMDD * Condition [Incongruent] | 0.92 | 0.25 – 1.60 | **0.008** |
| (ADHD * DMDD) * Condition [Congruent] | -0.28 | -1.10 – 0.54 | 0.505 |
| (ADHD * DMDD) * Condition [Incongruent] | -1.07 | -1.89 – -0.24 | **0.012** |
| **Random Effects** |  |  |  |
| $\sigma^2$ | 0.80 | | |
| $\tau_{00}$ Subject | 2.70 | | |
| ICC | 0.77 |  |  |
| N<sub>Subject</sub> | 187 |  |  |
| Observations | 561 |  |  |
| Marginal R<sup>2</sup> / Conditional R<sup>2</sup> | 0.373 / 0.857 |  |  |

a. References levels for factor contrasts are ADHD is absent, DMDD is absent, and Flanker condition is “Neutral.”

**Figure 3.**
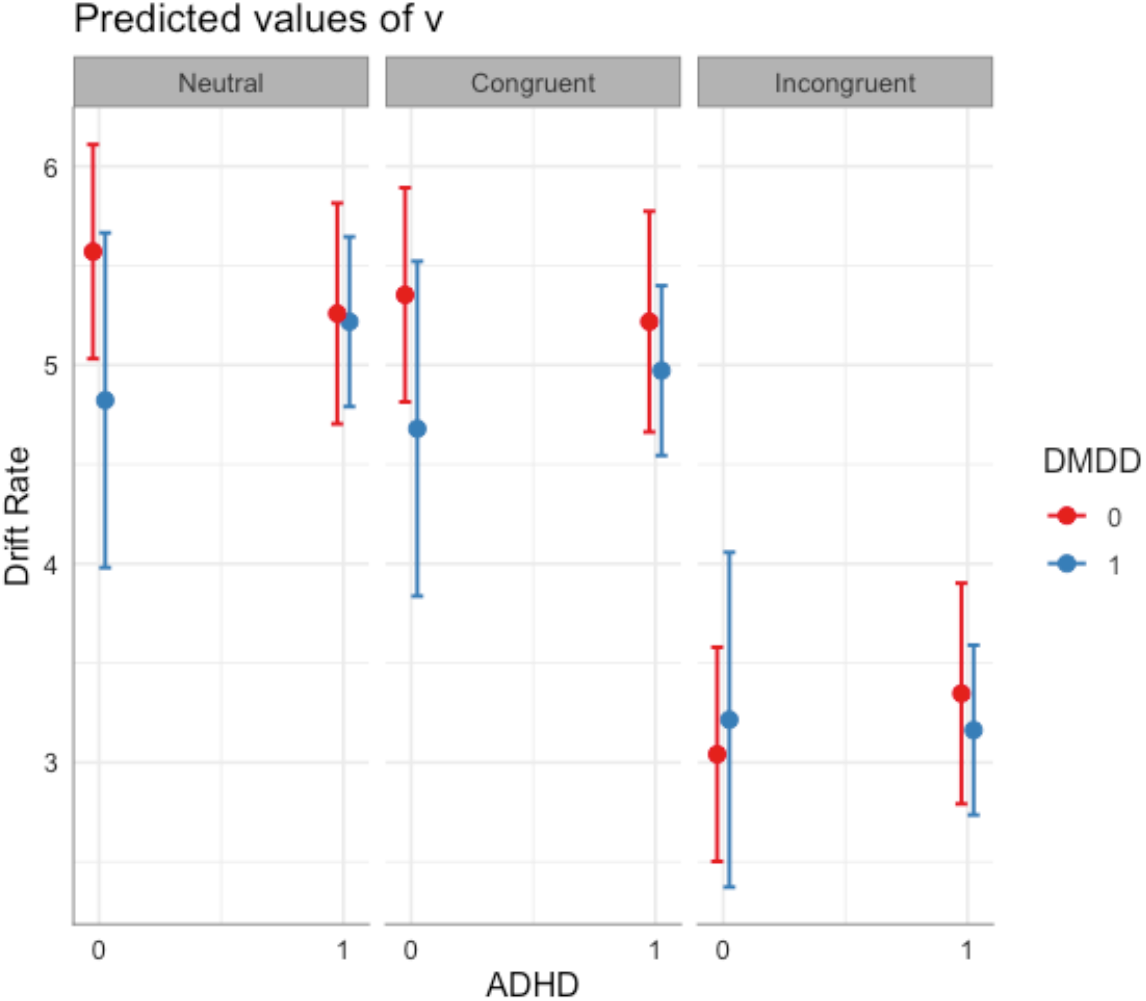
Effects of diagnostic status and task condition on drift rate *v*, adjusted for FSIQ and Age. For diagnostic codings, 1=Present and 0=Absent. Drift rate is lower in the incongruent condition relative to both neutral and congruent conditions. Differences between those with ADHD or DMDD are evident in the neutral and congruent condition. Error bars reflect 95% CI.

**Figure 4.**
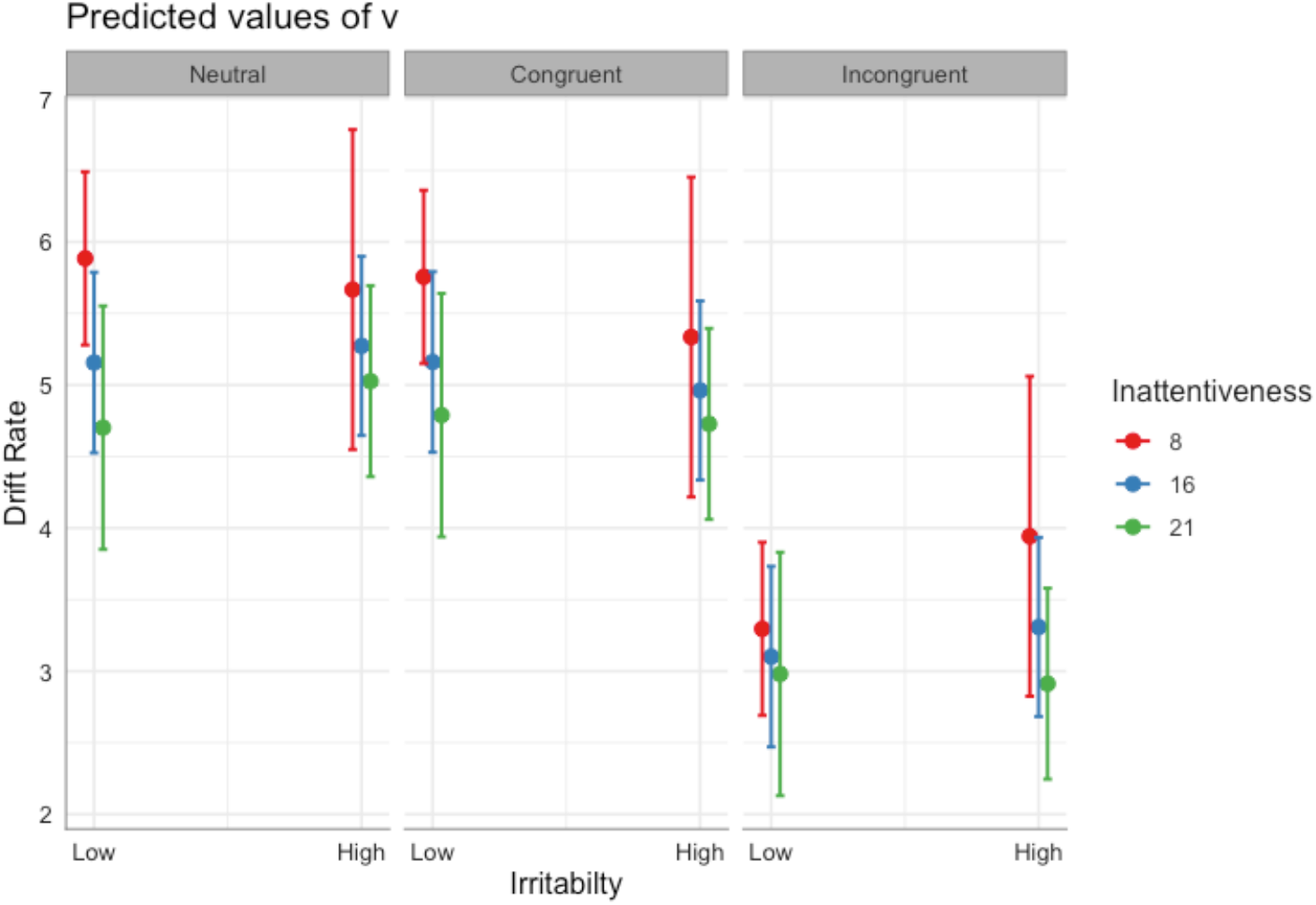
Effects of dimensionally-assessed irritability and inattention and task condition on drift rate *v*, adjusted for FSIQ and Age. Error bars reflect 95% CI. Increased CIs estimated for youth high in irritability and low in inattention are due to the moderate correlation among symptom domains.

### Symptom Associations

Additional analyses were conducted on symptoms in the smaller subsample (*n*=87). These analyses revealed interactions between both symptom dimensions and task condition on drift rate (see Table 3 for full model). Notably only inattentiveness was associated with reduced drift rate across all task conditions (b=−.09, *p*=.011, 95%CI[−.16, −.02]), while irritability was specific to task condition (b=1.64, *p*=.021, 95%CI[.26, −3.02]). However, similar to the categorical analysis, they interacted such that in combination they did not have purely additive effects. Notably, the two symptom dimensions interacted with task conditions such that irritability modulated the effects of inattention on drift rate by condition (b=−.10, *p*=.018, 95%CI[−.18, −.02]); see Figure 3).

**Table 3.**
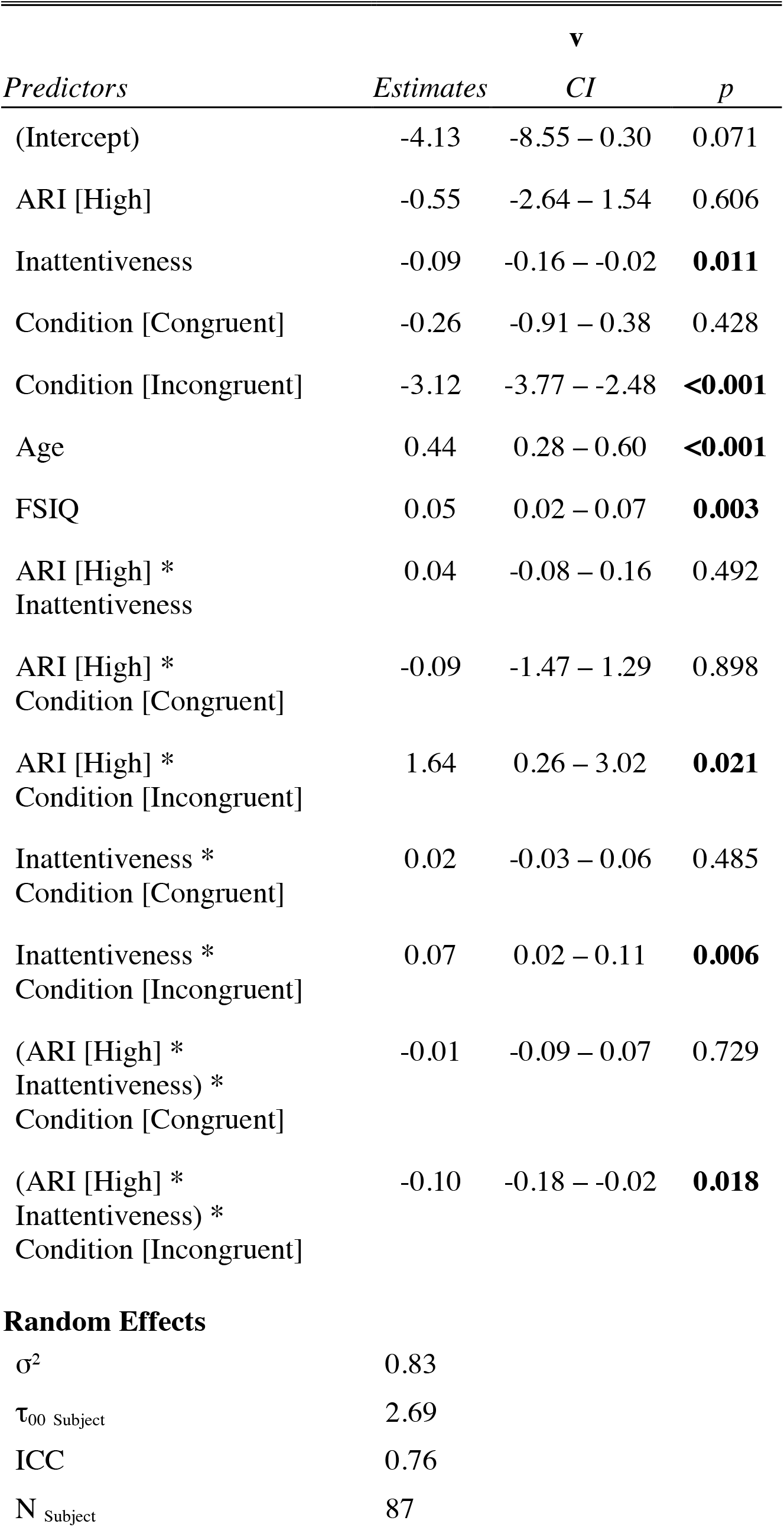
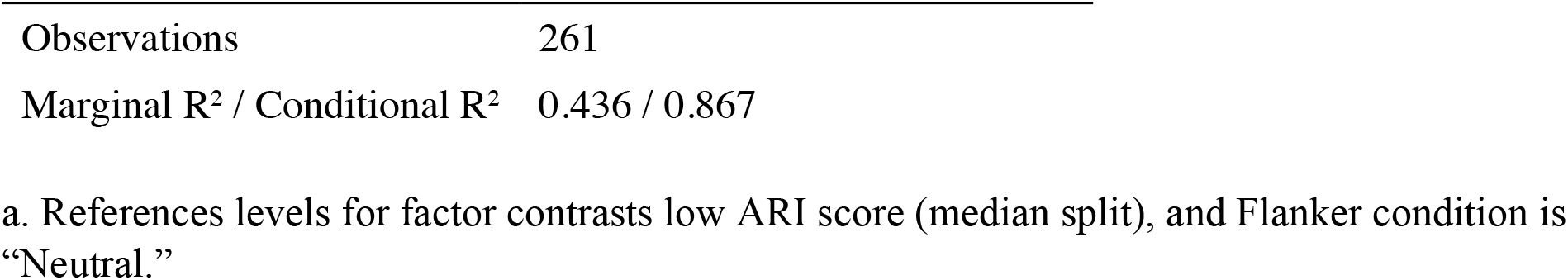
Mixed effects model regression table for drift rate predicted by symptom dimension.

| *Predictors* | *Estimates* | *v* |  |
| --- | --- | --- | --- |
|  |  | *CI* | *p* |
| (Intercept) | -4.13 | -8.55 – 0.30 | 0.071 |
| ARI [High] | -0.55 | -2.64 – 1.54 | 0.606 |
| Inattentiveness | -0.09 | -0.16 – -0.02 | **0.011** |
| Condition [Congruent] | -0.26 | -0.91 – 0.38 | 0.428 |
| Condition [Incongruent] | -3.12 | -3.77 – -2.48 | **<0.001** |
| Age | 0.44 | 0.28 – 0.60 | **<0.001** |
| FSIQ | 0.05 | 0.02 – 0.07 | **0.003** |
| ARI [High] *<br>Inattentiveness | 0.04 | -0.08 – 0.16 | 0.492 |
| ARI [High] *<br>Condition [Congruent] | -0.09 | -1.47 – 1.29 | 0.898 |
| ARI [High] *<br>Condition [Incongruent] | 1.64 | 0.26 – 3.02 | **0.021** |
| Inattentiveness *<br>Condition [Congruent] | 0.02 | -0.03 – 0.06 | 0.485 |
| Inattentiveness *<br>Condition [Incongruent] | 0.07 | 0.02 – 0.11 | **0.006** |
| (ARI [High] *<br>Inattentiveness) *<br>Condition [Congruent] | -0.01 | -0.09 – 0.07 | 0.729 |
| (ARI [High] *<br>Inattentiveness) *<br>Condition [Incongruent] | -0.10 | -0.18 – -0.02 | **0.018** |
| **Random Effects** |  |  |  |
| $\sigma^2$ | 0.83 | | |
| $\tau_{00}$ Subject | 2.69 | | |
| ICC | 0.76 |  |  |
| $N$ Subject | 87 | | |

|  |  |
| --- | --- |
| Observations | 261 |
| Marginal $R^2$ / Conditional $R^2$ | 0.436 / 0.867 |

a. References levels for factor contrasts low ARI score (median split), and Flanker condition is “Neutral.”

In the hyperactivity model, hyperactivity was associated with a reduced drift rate (b=−.09, *p*=.038, 95%CI[−.18, −.01]) similar to the pattern of inattentive symptoms, above, but no other main or interactive effects of symptoms were evident (see Supplementary Table S3 for full model).

For all linear models, all predictors had a corrected generalized variance inflation factor <5, suggesting acceptable levels of multicollinearity for analysis.

## Discussion

Here, we conduct the first examination of reaction time variability, via the DDM, in youth with severe irritability accounting for ADHD symptoms. We demonstrate common and distinct associations with drift rate, ADHD, DMDD, and their core symptoms that vary by attentional demands. Consistent with our hypotheses, drift rate, or processing efficiency, decreased by having ADHD or DMDD or being confronted with distractors. Specifically, increases in processing efficiency for non-conflict conditions were larger in youth without psychopathology relative to patients. Analyses of continuous measures in a smaller subsample revealed interactive effects between irritability and inattention on drift rate. Effects appeared particularly pronounced when switching from continuous performance demands to conflict processing (incongruent condition) settings.

As expected, the strong correlations occurred between response variability and the drift rate parameter; lower drift rates will result in a wider distribution of reactions times (Ratcliff & McKoon, 2008; Wagenmakers, van der Maas, & Grasman, 2007). This suggests that the increased variability seen in sustained attention tasks in youth with ADHD and DMDD can be partially explained by less efficient processing. Hence, the computational modeling perspective adds to our understanding of the behavioral dysfunction by bringing interpretational clarity to decision computation impairments observed in ADHD and DMDD. In the case of ADHD, DDM-measured cognitive processing inefficiency support hypotheses of neural processing inefficiency (i.e., lower signal to noise ratio in decision-making networks, Huang-Pollock et al., 2017). Empirically, efforts have been made to combine neural and computational approaches to map neural signals to specific computations captured in latent parameters (Turner, Forstmann, Love, Palmeri, & Van Maanen, 2017) or even undertake neutrally informed modeling (O’Connell, Shadlen, Wong-Lin, & Kelly, 2018).

Though much has been written about the involvement of sustained attention on response time variability in ADHD (Huang-Pollock et al., 2012; Karalunas et al., 2012), caution is warranted when interpreting cognitive control functions underlying interindividual differences in reaction time variance (Huang-Pollock et al., 2017). Attentional demands reflect cognitive load influences on processing efficiency (White et al., 2011). Notably, we find associations with diagnostic status were modulated by attentional demands, with the most pronounced associations in conditions of relatively low attentional demand. This may reflect a floor effect on drift rate, where high cognitive load suppresses drift rate to the extent that it obscures symptom associations. On the other hand, it may reflect symptom-related issues with effectively recruiting cognitive resources in conditions which require lower cognitive resources, e.g. cognitive flexibility.

The only previous fMRI study (Pagliaccio et al., 2017) to examine sustained attention in youth with ADHD and DMDD found blunting in parietal attention networks among both patients with ADHD and DMDD associated with longer trial-wise reaction times. Additionally, the study found DMDD-specific increases in pre-stimulus activation associated with longer trial-wise reaction times in several frontal and parietal regions. Converging with the current findings, this evidence suggests that cognitive control impairments are not specific to ADHD, but also link to DMDD, and perhaps to chronic irritability. Decomposing the cognitive processes of sustained attention reflected in reaction time on the neural level would be a natural next step to aid interpretation of neural findings.

### Limitations and future directions

There are several limitations to consider when interpreting the findings. First, these results may only be generalized to populations able to learn and complete the Flanker task. A number of youth (∼15%) were unable to train to adequate task performance and were not invited to complete the task. Second, with a correlation of *r*=.49, *p*<.001, there is significant overlap between the constructs of inattention and irritability, and issues of multicollinearity arise, biasing toward type II error. However, examinations of variance inflation factor found these to within generally acceptable limits (<5) for all terms in all models. Though *fast-dm* is well validated, it only allows for a constant drift rate. A drift rate that varies with time might better capture early attentional dynamics in the flanker, which are especially prominent in the incongruent condition (White et al. 2011). Examining these variables dimensionally enabled us to explore more fine-grained interactions among symptom domains, as opposed to categorical diagnoses. Such data was available only for a subsample for the current report. However, the current diagnostic comparisons are helpful. They were determined by semi-structured interview including multiple sources of information and arrived at by consensus among experts. They reflect differences in attention/hyperactivity and irritability on which ADHD and DMDD are solely based. Finally, they are highly relevant for clinical practice and directly inform clinicians on the specificity of response time variability to ADHD. Future studies may leverage dimensional data via latent class modeling of clinical variables to examine how response time variability and processing efficiency track with individual differences in irritability and ADHD symptoms. An interesting future direction is examining cognitive control in an affective, rather than a ‘cold’ cognitive context. Cognitive control processes contribute to the regulation of affective states, i.e. emotional regulation (Ochsner & Gross, 2005). Current models of chronic, severe irritability posit impairments in cognitive control as a potential mediator for experiences of frustration and behavioral manifestations of irritability, i.e., temper outbursts (Kircanski et al., 2019). Hence, ‘cold’ cognitive control differences observed in the current study may be magnified in cognitive control tasks where goal conflicts evoke negative affect.

## Conclusion

Applying a computational modeling approach, we can map increased variability in reaction times in ADHD and DMDD onto difficulties in basic processing of stimuli under different attentional demands. Our results suggest that attentional impairments are not specific to ADHD; rather they may represent shared psychopathology between ADHD and DMDD.

## Data Availability

Data will be made available in compliance with NIH/NIMH guidelines.

## Notes

### Competing Interest Statement

The authors have declared no competing interest.

### Funding Statement

This research was supported by the Intramural Research Program of the NIMH, National Institutes of Health (NIH; ZIAMH002786), and was conducted under NIH Clinical Study Protocols 00-M-0198 and 02-M-0021 (ClinicalTrials.gov ID: NCT00006177 and NCT00025935). JS was supported by a grant from the National Institutes of Health, National Institute of Mental Health, K23MH113731 and the Pediatric Mental Health Institute at Children’s Hospital Colorado and the Division of Child and Adolescent Psychiatry, Department of Psychiatry, University of Colorado School of Medicine. The funding source was not involved in study design; the collection, analysis and interpretation of data; writing of the report; and the decision to submit the article for publication.

